# Genomic epidemiology of early SARS-CoV-2 transmission dynamics in Gujarat, India

**DOI:** 10.1101/2021.08.31.21262680

**Authors:** Jayna Raghwani, Louis du Plessis, John T McCrone, Sarah C. Hill, Kris V. Parag, Julien Thézé, Dinesh Kumar, Apurva Puvar, Ramesh Pandit, Oliver G. Pybus, Guillaume Fournié, Madhvi Joshi, Chaitanya Joshi

## Abstract

Genomic surveillance of SARS-CoV-2 has played a decisive role in understanding the transmission and evolution of the virus during its emergence and continued circulation. However, limited genomic sampling in many high-incidence countries has impeded detailed studies of SARS-CoV-2 genomic epidemiology. Consequently, critical questions remain about the generation and global distribution of virus genetic diversity. To address this gap, we investigated SARS-CoV-2 transmission dynamics in Gujarat, India, during its first epidemic wave and shed light on virus’ spread in one of the pandemic’s hardest-hit regions. By integrating regional case data and 434 whole virus genome sequences sampled across 20 districts from March to July 2020, we reconstructed the epidemic dynamics and spatial spread of SARS-CoV-2 in Gujarat, India. Our findings revealed that global and regional connectivity, along with population density, were significant drivers of the Gujarat SARS-CoV-2 outbreak. The three most populous districts in Gujarat accounted ∼84% of total cases during the first wave. Moreover, we detected over 100 virus lineage introductions, which were primarily associated with international travel. Within Gujarat, virus dissemination occurred predominantly from densely populated regions to geographically proximate locations with low-population density. Our findings suggest SARS-CoV-2 transmission follows a gravity model in India, with urban centres contributing disproportionately to onward virus spread.

## Introduction

Global genomic surveillance of SARS-CoV-2 has provided key insights into virus dissemination and evolution at local, national, and international scales. Detailed analysis of the UK epidemic during the first wave^1^ revealed significant heterogeneity in the size and duration of different SARS-CoV-2 lineages, which was driven by high extinction rates and rapid fluctuations in virus importation. This pattern has been repeatedly observed globally; shifts in human mobility drastically alter the source of virus importations and only a small proportion of importations lead to sustained community transmission^2–7^. Virus importations can sometimes be sufficiently intense to have a significant effect on epidemic dynamics (e.g., super-seeding of alpha variant in the UK^8^). Genomic surveillance has been fundamental in tracking these dynamics, including detecting the emergence and spread of novel variants that alter risk to public health^9–11^.

Gujarat is the fifth largest and ninth most populous of India’s 28 states and has over 60 million inhabitants. Like the rest of India, most people live in rural areas (57.4%), although the proportion of people living in cities has been increasing in past decades due to urbanisation (from 37.4% in 2001 to 42.6% in 2011)^12^. Gujarat shares an international border with Pakistan, as well as Indian state borders with Rajasthan and Madhya Pradesh to the northeast and east, and Maharashtra and the Union Territories of Daman, Diu and Nagar Haveli, to the south. The region has two international airports in Ahmedabad and Surat, which served ∼11 and ∼1.5 million passengers^13^, respectively, between April 2019 and March 2020 (Fig 1A). While the first COVID-19 cases in India were reported on January 30, 2020, and linked to travel from Wuhan, China, the first cases in Gujarat were identified on March 19, 2020 and were associated with travel from Saudi Arabia and the UK. A few days later, on March 24, 2020, a nationwide lockdown across India was announced. Nevertheless, in Gujarat, special transportation (trains and buses) was arranged between March and May 2020 to allow ∼1.8 million stranded migrant workers to return to their home states (i.e., outside of Gujarat) or districts within Gujarat. The majority of specially organised trains originated from Gujarat (26%), which has one of the largest populations of migrant workers in India^14^.

**Figure 1:**
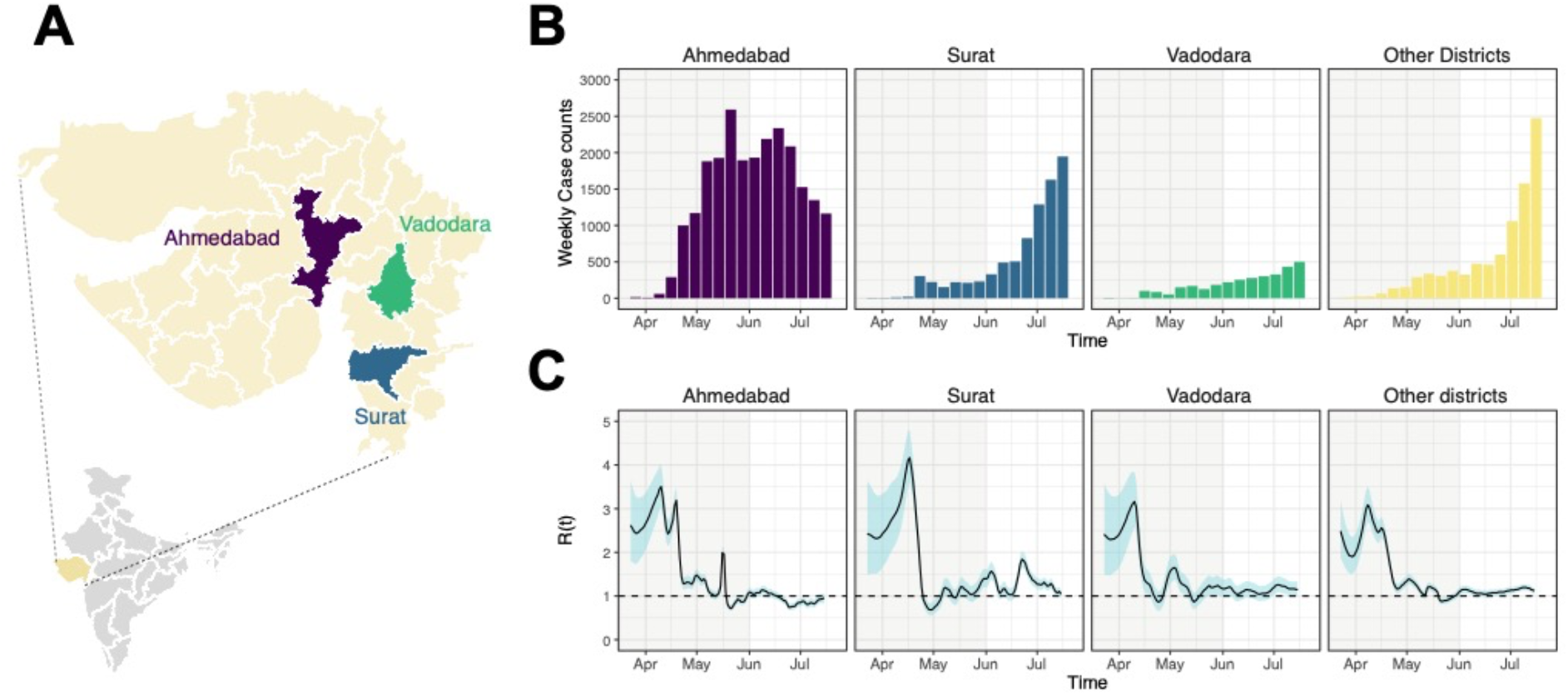
SARS-CoV-2 epidemiology during the first wave in Gujarat, India. (A) Map of Gujarat highlighting the three most populous districts (Ahmedabad = Purple, Surat = Blue, Vadodara = Green, all other districts = Yellow). (B) Weekly counts of newly-reported SARS-CoV-2 cases for Ahmedabad, Surat, Vadodara, and other districts from April to July 2020. (C) Estimates of the epidemic instantaneous reproduction number R(t) for four locations specified in panel B. The black line corresponds to the median estimate, while the shaded region corresponds to 95% equal tailed Bayesian credible intervals. The shaded region indicates the period of national lockdown.

In this study, we investigate the introduction and transmission of SARS-CoV-2 in Gujarat during the first wave of the COVID-19 epidemic in India, using a combination of epidemiological and genomic data. During the period April to July 2020, we used a multiplex PCR amplification and IonTorrent sequencing to generate 434 SARS-CoV-2 genome sequences. Through our analyses, we characterise the epidemiological and lineage dynamics of SARS-CoV-2, evaluate key drivers of virus importation and transmission, and assess whether significant changes in human mobility shaped virus transmission dynamics in Gujarat.

## Results

Analysis of the epidemic’s instantaneous reproduction number over time (R(t)) indicated that the three most populous districts (Fig 1A; Ahmedabad, Surat, and Vadodara) experienced rapid epidemic growth between late March to mid-April (Figure 1C). However, after this initial period of growth, a sharp decrease in R(t) was observed in all three major cities (Fig 1C). The initial growth period between March and April occurred during the period of national lockdown. This might be a consequence of a time lag between the number of reported cases and the epidemiological effect of the lockdown (which began on March 24, 2020) or might reflect considerable population movements during the early phase of the lockdown, for example due to individuals returning to their home states or districts. After the lockdown ended on June 1, 2020 (Fig 1; end of shaded region), Ahmedabad was characterised by slower epidemic growth, indicating that restriction in human mobility had a notable impact on disrupting chains of transmission. In contrast, in other districts, including Surat and Vadodara, from first week in June, the number of cases started to rapidly increase. Despite Ahmedabad, Surat, and Vadodara recording their first cases on 19-20 March, Ahmedabad accounted for majority of confirmed cases during the first wave (Fig 1B). This pattern is consistent with faster epidemic progression in Ahmedabad, possibly driven by higher frequency of virus importation compared to other districts, which have comparatively weaker international links due to lack of airports or significantly lower passenger flow, and differences in population density and population connectedness.

To better understand the source of virus importations in Gujarat, we characterised the lineage dynamics of SARS-CoV-2 in the region using phylogenetic analyses. Specifically, we combined 434 genome sequences generated from Gujarat with a representative subset of global genetic diversity of SARS-CoV-2 sampled over a similar period. We detected 39 (95% HPD: 33-44) distinct Gujarat transmission lineages (defined as comprising two or more Gujarat genomes that belong to an ancestral lineage that originates from outside of Gujarat), comprising 360 sequences, and 74 (95% HPD: 63-83) singletons (genomes that could not be allocated to a Gujarat transmission lineage) across 2000 posterior trees (Fig 2A). In total, 113 virus importations were identified into Gujarat during the first wave. Consistent with previous analyses ^1^, larger transmission lineages were associated with longer duration times (the time between the oldest and most recently sampled genome within a transmission lineage), while smaller transmission lineages were associated with shorter duration times (Fig 2A). This observation is further corroborated by the strong negative exponential relationship between transmission lineage size and the time of the most recent common ancestor (TMRCA), as shown in Fig 2B. The mean TMRCA of transmission lineages was on April 17, 2020 (SD = 35.8 days), with 75% of transmission lineages having a TMRCA between 28 March and May 7, 2020, which coincides with the period when the epidemic was growing exponentially.

**Figure 2:**
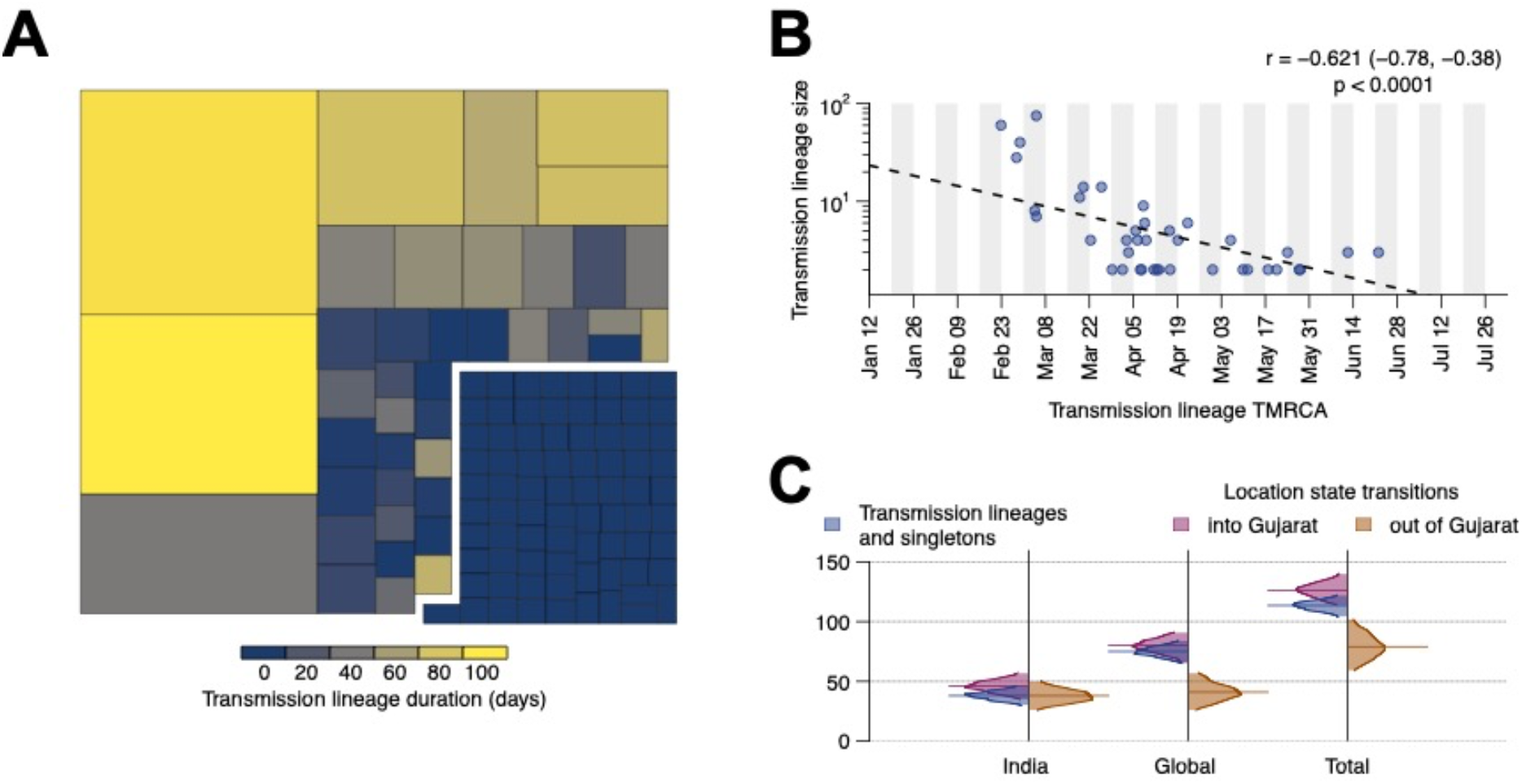
Size, duration, and importations of SARS-CoV-2 transmission lineages in Gujarat. A) Treemap summarising the 113 detected transmission lineages by size. Colours indicate the duration of persistence of the lineage and the areas indicate the size of the transmission lineages. Lineage duration corresponds to time between the lineage’s oldest and most recently sampled genomes. B) There is a strong log-linear relationship between size and mean TMRCA (time to most recent common ancestor) of each transmission lineages. C) Breakdown of virus importations into Gujarat from other lndian states or countries. The number of location state transitions were estimated using a robust counting approach ^15^ and a three-location discrete trait phylogeographic analysis.

To determine likely sources of virus importation into Gujarat, we performed a three-location discrete trait phylogeographic analysis (with location states of Gujarat, India (excluding Gujarat), and global). Interestingly, despite sharing a border with other states with high disease burden (e.g., Maharashtra), our results suggest that virus importations into Gujarat have been driven primarily by international travel. However, the relatively low frequency of virus genome sampling across India is likely to mask importations from other Indian states. Notably, over the study period, there were 3092 SARS-CoV-2 genomes available from India suitable for phylogenetic analysis (high-coverage and with complete temporal information), and only 930 were from neighbouring states, predominantly from Maharashtra.

Next, to evaluate the spatial dynamics of SARS-CoV-2 within Gujarat, we undertook a more detailed phylogeographic analysis to evaluate predictors of virus lineage dissemination using a generalised linear model. To ensure the results reflect the specific dynamics of SARS-CoV-2 in Gujarat, we included only those sequences from the seven largest Gujarat transmission lineages (Figure 3B), which contained between 11 to 75 sequences. All seven lineages were associated with virus importation from outside of India with the mean TMRCA occurring close to the start or before the national lockdown. Four lineages were mainly comprised of genome sequences sampled from Ahmedabad, two were associated with Vadodara and/or Surat, and one was associated with Aravalli district. The main predictors we tested were case counts, population size, and population density at both origin and destination locations, and the road distance between districts (Figure 3A). Overall, our findings indicate that viral lineages moved (i) more intensely between districts that were geographically closer (BF > 100; Fig 3C) and (ii) predominantly from districts with higher population density to districts with lower population density (BF = 40; Fig 3C).

**Figure 3.**
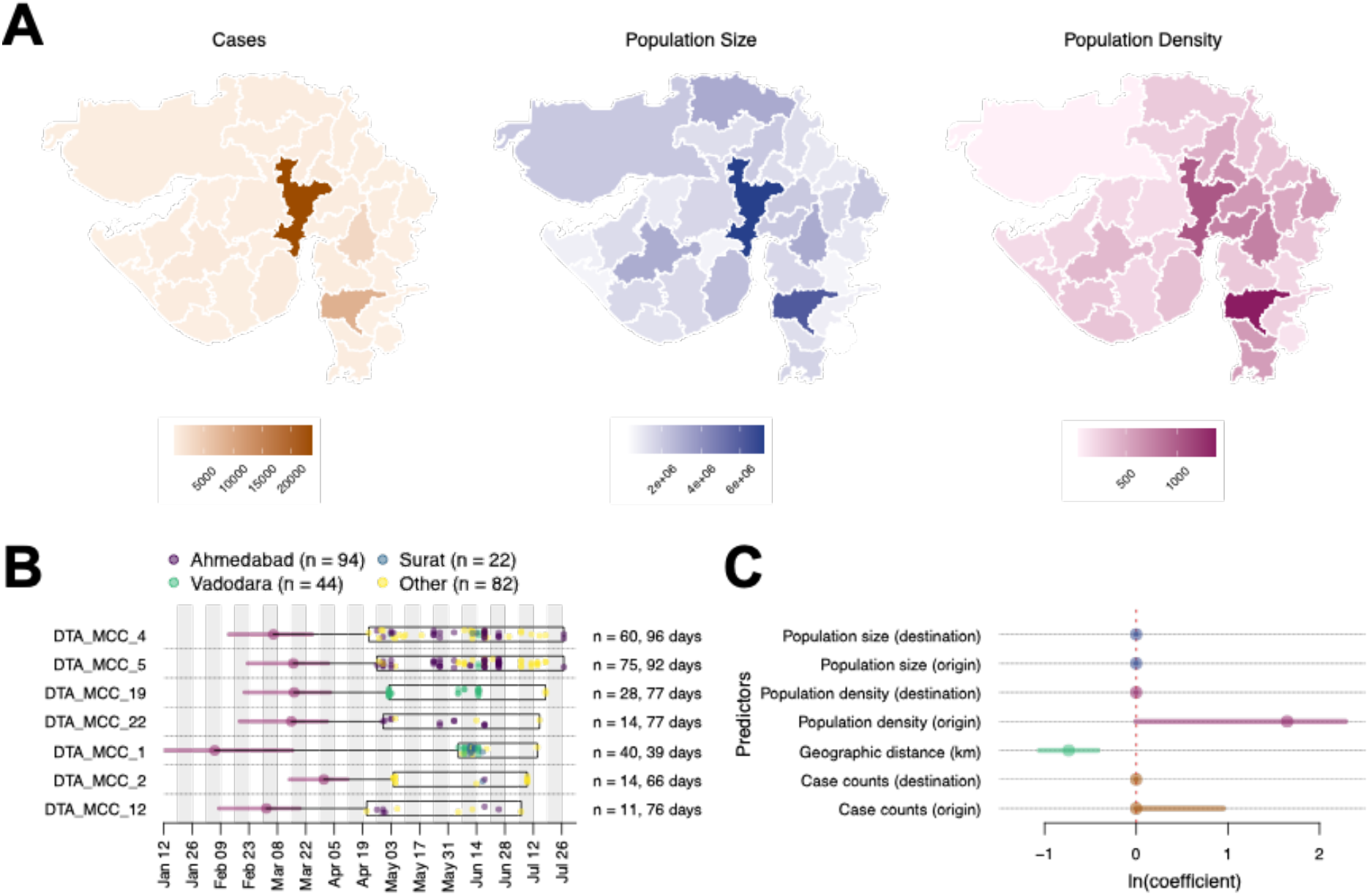
Determinants of SARS-CoV-2 lineage spread within Gujarat: (A) Choropleth maps of key predictors (cases, population size, and population density) that were evaluated in the phylogeographic generalised linear model analysis along with geographic distance. (B) TMRCA and sample distribution of the seven largest transmission lineages. For each lineage, circles correspond to the estimated lineage TMRCA, and horizontal bars indicate the 95% HPD interval of the TMRCA. The box shows the date range of the samples from Gujarat for each lineage. The total number of samples and duration of each lineage are shown on the right-hand side. C) Predictors of SARS-CoV-2 lineage movement within Gujarat based on 20 sampled districts. The contribution of each predictor is represented by the mean coefficient value (points) and 95% HPD interval (horizontal bars).

Together with the epidemiological data (Fig 1C), these results suggest that viral lineage movement was concentrated in exports from urban centres, which had higher caseloads and population densities, to nearby districts – initially from Ahmedabad but at later stages also from Surat and Vadodara. These movements contributed to the spread of SARS-CoV-2 within Gujarat.

Although the global phylogeographic analysis suggests that importations of transmission lineages to Gujarat were mainly associated with international travel, phylogenetic analysis of the two largest Gujarat transmission lineages (Fig 4) provides evidence that these lineages were subsequently exported globally and to other states in India. Clusters of (two or more) sequences from Canada, Oman, Karnataka, and Odisha were detected nested within DTA_MCC_4 (Fig 4A), suggesting that this transmission lineage may have seeded outbreaks in these locations. Furthermore, in DTA_MCC_5, we detected clusters of sequences from Bangladesh and Telangana, as well as singletons from the USA, UAE, Hong Kong, and France. A number of these non-Gujarat clusters and singletons are on long branches, which means that we cannot exclude the possibility of intermediate locations in viral exportation.

**Figure 4.**
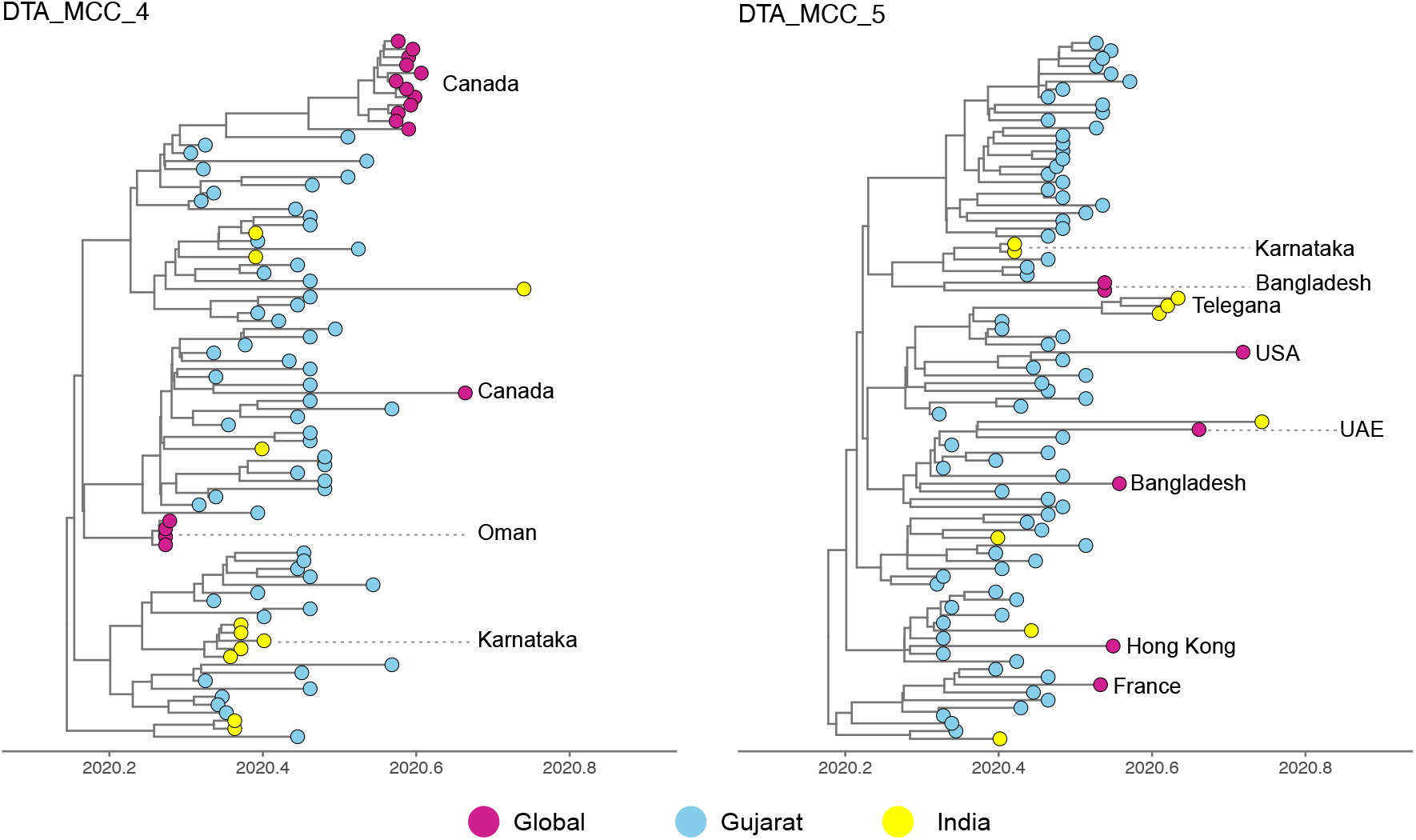
Virus exportation from Gujarat: Maximum clade credibility trees for the two largest transmission lineages identified in this study: (A) DTA_MCC_4 and (B) DTA_MCC_5.

## Discussion

Using epidemiological data and virus whole genome sequences, we investigated the spread, importations, and lineage dynamics of the first wave of SARS-CoV-2 in Gujarat, India. Initially, the epidemic was concentrated mainly in the most populous district, Ahmedabad, which accounted for more than half of reported cases. Epidemiological analysis revealed that R(t) decreased across all districts after the national lockdown was announced. However, despite the slowdown in epidemic growth, the number of cases continued to rise across Gujarat after restrictions were gradually lifted on May 31, 2020, particularly in other populous districts, such as Surat and Vadodara. This suggests that some transmission chains were able to persist during the lockdown, which then resumed circulation and growth after the lockdown ended. Phylogenetic analyses suggest that virus importations into Gujarat were comparatively higher than virus exportation from Gujarat, and that virus importations were predominantly associated with international travel. This pattern is consistent with the disproportionately higher case burden in Ahmedabad, which hosts the busiest airport in the state and a major destination for interstate movement in Gujarat.

While our findings suggest that the mass transportation of migrant workers within and from Gujarat were not a significant driver of virus transmission in Gujarat, the comparatively low genomic surveillance across India means that our study is likely to underestimate the number and rate of within-country virus movements. Nevertheless, our phylogeographic analysis identified geographic proximity between locations as a key driver of virus transmission in Gujarat, which was similarly observed for the alpha variant in the UK^8^. This suggests that disease control strategies should consider spatial context of SARS-CoV-2 spread, e.g., interventions should not only focus on populations with high disease prevalence but also be expanded to include geographically proximate populations to limit onward transmission. Particularly, more coordinated, responsive approaches at the local level could prevent travelling waves of infections without the need for national lockdowns^16^. This strategy would likely require an improved understanding of multi-scale mobility patterns across India and elsewhere to better respond to such epidemics^17^.

As noted in the UK, virus importation is expected to occur earlier than the estimated TMRCA of a transmission lineage (between 0 to 10 days) ^1^. However, due to a lack of detailed data on population movements into Gujarat, we were not able to estimate importation dates for the Gujarat transmission lineages. Nonetheless, the TMRCAs of transmission lineages serve as upper bounds on the importation events (i.e., importation must occur before the TMRCA). The earliest transmission lineages (based on TMRCA) were associated with importation from outside of India, although due to the relatively small number of genomes from Gujarat, it was not possible to evaluate changes in the dynamics of virus importation before and after the lockdown. However, like previous studies ^1,4^, transmission lineages with earlier TMRCAs tended to be larger and have longer duration times.

Our analyses indicates that the Gujarat epidemic during the first wave was associated with over 100 virus introductions. Given that this estimate was obtained from a relatively small number of sampled genomes, the true number is likely to be much larger. The faster progression of the epidemic in Ahmedabad compared to other districts strongly suggests this district was the epicentre of the first wave, likely enabled by a combination of higher inflow of international travellers, population density and connectedness to the rest of Gujarat. Notably, the focus of transmission started to shift to other districts, such as Surat and Vadodara, after July 2020, perhaps as result of regional differences in human mobility and behaviour (e.g., less movement or greater caution in Ahmedabad compared to other districts). However, due to the limited temporal range of the sampled genomes, we could not test this hypothesis phylogenetically. Nevertheless, rapid advancement of epidemics in urbanized regions which later move onto less populous regions has been commonly observed elsewhere^18–20^, including during the 2009 H1N1 influenza A pandemic^21^. Importantly, this strongly suggests that to reduce disease transmission, interventions should be implemented rapidly and robustly in major urban centres (e.g., as demonstrated in China in early 2020^22^).

Although our study period precedes the detection, emergence, and international spread of the delta variant of concern (Pango lineage B.1.617.2) in 2021, insights about SARS-CoV-2 transmission dynamics in Gujarat, India has implications for understanding how the delta variant arose and spread within India, and subsequently worldwide. Notably, we show the importance of international connectedness and intra-regional demographics in shaping virus lineage dynamics in India, and we highlight the limitations of investigating virus movement and origins due to the comparatively low genomic surveillance in this region. All of these factors will need to be taken into consideration when evaluating the emergence and epidemiology of the delta variant.

## Methods

### Sample collection, library preparation, sequencing and data analysis

Nasopharyngeal/oropharyngeal swabs from COVID-19 positive individuals were collected after obtaining informed consent and ethics approval. Samples were transported and processed for sequencing as previously described in ^23^. In brief, we used the Ion AmpliSeq SARS-CoV-2 research panel and Ion AmpliSeq™ Library Kit Plus for the library preparation. Sequencing was performed on Ion Torrent S5Plus system using a 530 chip with 400bp chemistry. We used a reference-based genome assembly, as described in^23^, to obtain whole genome sequences. In brief, we used PRINSEQ-lite v.0.20.4^24^ for trimming and quality filtering. High quality reads were mapped against a SARS-CoV-2 reference genome (GenBank accession number NC_045512) using CLC Genomics Workbench V 12.0 to obtain consensus genomes.

### Epidemiological analysis

COVID19 case data was obtained from a crowdsourced initiative, hosted at https://www.covid19india.org/ and https://api.covid19india.org/. This data has been curated by volunteers from different data sources such as State press bulletins and the Ministry of Health & Family Welfare of the Government of Gujarat. The source code is available in the GitHub repository at https://github.com/covid19india/api.

We estimated the instantaneous effective reproduction number at time t, R(t) (median and 95% equal tailed Bayesian credible intervals shown in Fig 1), using the EpiFilter method^25^. This approach applies optimal recursive smoothing techniques to minimise the mean squared error in inferring R(t) from the incidence of cases under a renewal transmission model. For all analyses we assumed that the SARS-CoV-2 generation time distribution is well approximated by the serial interval distribution from ^26^ and we apply a weekly averaging filter to reduce weekend effects and inconsistent reporting, which likely corrupts the incidence time series.

### Identification of transmission lineages

Based on the lineages detected in the Gujarat SARS-CoV-2 dataset, we collated a representative global dataset comprising 10,000 genome sequences sampled evenly by week and country. To maximize our power to detect within-India dissemination, we added all Indian sequences available on GISAID as of November 5, 2020 to this dataset, for a final data set size of 12,180 sequences (including samples from Gujarat). We followed a similar pipeline to that outlined in du Plessis et al and used ThorneyBeast (https://beast.community/thorney_beast) to estimate the posterior molecular clock tree. Briefly, a maximum likelihood tree was estimated with iqtree, using a Juke-Cantor substitution model with Wuhan/WH04/2020 as on outgroup. Branch lengths in this tree were scaled and rounded to represent the expected number of mutations. We then estimated the posterior tree distribution under a Skygrid coalescent prior^27^, and a strict molecular clock model with a fixed rate (0.00075 subs/site/year). We executed 10 chains of 400 million steps, logging every 7.2 million steps with the first 40 million steps removed as burnin.

We used a three-state asymmetric discrete trait analysis (DTA) model implemented in BEAST v1.10^28^ to infer the ancestral node locations (Gujarat, India, or global) using an empirical tree distribution comprising 500 time-calibrated trees sampled from the posterior tree distributions estimated above.

To identify transmission lineages, we followed the methodology described in du Plessis et al. Briefly, we performed a depth-first search from any movement from either global or India to Gujarat. Using this approach, we identified seven transmission lineages with >10 sequences, which were selected for further analysis.

### Drivers of virus transmission within Gujarat

To investigate the drivers of virus transmission, we focused on the seven largest transmission lineages identified in the global dataset, as described above. Each dataset ranged from 11 to 75 sequences. Data on key predictors (population size and density, number of cases, geographic distance) were collated at the district level. For each transmission lineage dataset, we employed a separate exponential coalescent prior, while sharing a SRD06 substitutional model^29^, a strict molecular clock model with a fixed rate (0.001 subs/site/year), and generalised linear phylogeographic model^30^. Eight chains of 50 million steps (logged every 5000 steps) were executed. The chains were combined and thinned by a factor of 10 after discarding the first 10% as burnin.

## Data Availability

All data used in this study were collated from public databases and therefore are freely available. Regional case data, SARS-CoV-2 genome sequences and analysis files used in this study have been made available via https://github.com/jnarag/covid-gujarat.

## ACKNOWLEDGEMENTS

The sequencing work at GBRC was supported by Department of Science & Technology, Government of Gujarat. JR, GF, DK, AP, RP, OP, MJ, and CJ were supported by the UKRI GCRF One Health Poultry Hub (Grant No. BB/S011269/1), one of twelve interdisciplinary research hubs funded under the UK government’s Grand Challenge Research Fund Interdisciplinary Research Hub initiative. SCH was supported by the Wellcome Trust [220414/Z/20/Z]. This research was funded in whole, or in part, by the Wellcome Trust [Grant number 220414/Z/20/Z]. For the purpose of open access, the author has applied a CC BY public copyright licence to any Author Accepted Manuscript version arising from this submission.

